# Identifying key factors in predicting Chikungunya and Zika transmission in French Polynesia: a data-driven mathematical model

**DOI:** 10.1101/2023.05.11.23289868

**Authors:** Zhiyuan Yu, Xi Huo, Peter J. Thomas, Qimin Huang

## Abstract

**Background:** Chikungunya and Zika are both arboviruses transmitted through the *Aedes* mosquitoes, which are ectothermic, leading to seasonal outbreak patterns of virus infections in the human population. Mathematical models linked with mosquito trap data, human case data, or both, have proven to be powerful tools for understanding the transmission dynamics of arboviral diseases. However, while predictive models should consider a variety of features in the environment, vectors, and hosts, it is not clear which aspects are essential to assist with short-term forecasting.

**Methodology:** We consider four simple models with various assumptions, including mosquito dynamics, temperature impacts, or both, and apply each model to forecast the Chikungunya and Zika outbreaks of nine different regions in French Polynesia. We use standard statistical criteria to compare the accuracy of each model in predicting the magnitude of the outbreak to select the most appropriate model to use as an alert system for arbovirus infections. Moreover, by calibrating our “best model”, we estimate biologically meaningful parameter values to explore the commonality and difference between Chikungunya and Zika epidemics.

**Conclusions:** We show that incorporating the mosquito population dynamics in the arbovirus transmission model is essential for accurate arbovirus case prediction. In addition, such enhancement in the accuracy of prediction is more obvious for the Chikungunya data than the Zika data, suggesting that mosquito dynamics play a more important role in Chikungunya transmission than Zika transmission. In contrast, incorporating the effects of temperature may not be necessary for past outbreaks in French Polynesia. With the well-calibrated model, we observe that the Chikungunya virus has similar but slightly higher transmissibility than the Zika virus in most regions. The best-fit parameters for the mosquito model suggest that Chikungunya has a relatively longer mosquito infectious period and a higher mosquito-to-human transmission rate. Further, our findings suggest that universal vector control plans will help prevent future Zika outbreaks. In contrast, targeted control plans focusing on specific mosquito species could benefit the prevention of Chikungunya outbreaks.

## 1 Introduction

Arbovirus refers to the type of virus spread to people by the bite of blood-feeding insects such as mosquitoes and ticks [1]. Chikungunya fever and Zika fever are diseases caused by arboviral infections and such arboviruses are transmitted via mosquitoes of the *Aedes* genus [2]. In recent years, these two diseases have spread quickly across the world, and several large outbreaks have occurred in the regions of India, the Pacific Ocean, and America [3]. In addition, arboviruses are likely to expand to other countries via the travels of asymptomatic individuals. Predictive and inferential models of arboviral disease outbreaks can assist public health authorities in raising awareness in local communities, making travel recommendations, and conducting proper vector control interventions.

Mathematical modeling for vector-borne diseases can be traced back to 1911 when Sir Ronald Ross proposed several difference equations to describe the transmission of Malaria between people and mosquitos [4]. In 1927, Kermack and McKendrick extended Ross’s equations into continuous-time differential equations and introduced the mass-action transmission term into the equations. Kermack and McKendrick’s pioneering compartmental SIR model later became the fundamental structure of the modern epidemic models [5–7]. Ross’s work was further refined by Macdonald in 1952 and led to the well-known Ross-Macdonald model [8]. In 1979, Anderson and May refined the Kermack-McKendrick model by introducing birth and death, which is more realistic and is therefore widely used [9]. Today, modeling approaches for infectious diseases can be classified into five categories: compartmental model, spatial model, network model, and individual-based model [10]. In the vector-borne compartmental models, the human population and the vector population are each split into compartments such as susceptible, infectious, etc. For instance, Funk et al. (2016) developed an ordinary differential equation model, a variant of the Ross-Macdonald model, to predict and compare the transmissions of Dengue and Zika outbreaks [11]. In their model, the human population is separated into susceptible, exposed, infectious, and recovered, and the mosquito population is separated into susceptible, incubating, and infectious. Bonyah and Okosun (2016) developed an ordinary differential equation model with optimal control to predict the transmission of Zika and evaluate the effects of treatment and insecticide [12]. Recently, Chen and Huo (2023) developed mathematical models to investigate the connections between the environmental factors and the *Aedes aegypti* population dynamics [13]. Despite the differences in the formulation, one of the most important parameters that can be measured by the compartmental epidemic models is the basic reproductive number (*R*_0_), which is defined as the average number of secondary infections produced by one infectious individual at the beginning of the outbreak [10]. *R*_0_ is an indicator of the transmissibility of a disease, and it can vary with place and time for the same disease. Other modeling approaches for vector-borne diseases include spatial modeling that models the transmission with certain transmission kernels [14], network modeling that uses a graph to model the interactions between people and disease-transmitting vectors [15], and individual-based modeling that can incorporate complex individual-level behaviors, interactions, and characteristics [16].

While complex models can capture a lot more details and achieve much better training and testing accuracy with careful tuning, simple models are still competitive for their interpretability and robustness, which sometimes make them better inferential tools than complex models. Thus, we aim to develop a simple data-driven framework to predict the transmission of vector-borne disease and identify important factors in vector-borne transmission, and we derived several biological meaningful parameters from the framework. An ideal inferential framework should contain as few parameters as possible such that the parameter estimates will be consistent across different sets of training data obtained from the same or similar biological settings. Also, the model should be complex enough to reflect any meaningful biological variations.

Uninfected mosquitoes obtain the virus by biting an infected person. Then the virus-carrying mosquitoes can cause new human infections if they bite susceptible people afterward. Mosquitoes are ectothermic, meaning their reproduction, development, feeding, and survival rates are sensitive to the external temperature. Therefore, researchers believe it is essential to consider the abundance of mosquitoes in the area, the mosquito infection dynamics, and their variation based on the local temperature fluctuation when developing compartmental models of arbovirus outbreaks. For example, most previous studies that revisit past arbovirus outbreaks have considered both mosquito population dynamics and temperature fluctuations [17–23]. On the other hand, arbovirus outbreaks often occur in warm weather and in sub-tropical and tropical areas, with limited temperature variation during the outbreak periods. Thus some models targeting ongoing outbreaks may interpret the case data without incorporating the temperature data [24] or the mosquito population dynamics.

This study investigates the roles of two factors in arbovirus outbreaks: mosquito population dynamics and local temperature. To do so, we consider four compartmental models with and without these two features. Outbreak data for Chikungunya and Zika fevers, together with local temperature, are adopted from a prior study [3] in nine different regions of French Polynesia. Specifically, we use each model as a forecasting tool to inform weekly case reports and evaluate the quality of prediction statistically, in order to select the optimal model in terms of simplicity and accuracy.

Once we establish the optimal model, we use it to address several biological questions about the outbreak data shown in Figure 1. Firstly, although the primary mosquito species that transmit Zika and Chikungunya are the same, most regions experience outbreaks on different scales. Moreover, we seek a qualitative explanation of the factors that caused different outbreak sizes of both diseases in the same area. Secondly, we observe that all nine regions experienced similar Zika outbreak sizes; we explore whether this commonality indicates shared disease transmission features across different areas. Finally, we offer an explanation for the occurence of the largest Chikungunya outbreak, in the Marquesas Islands, compared to other regions.

**Figure 1:**
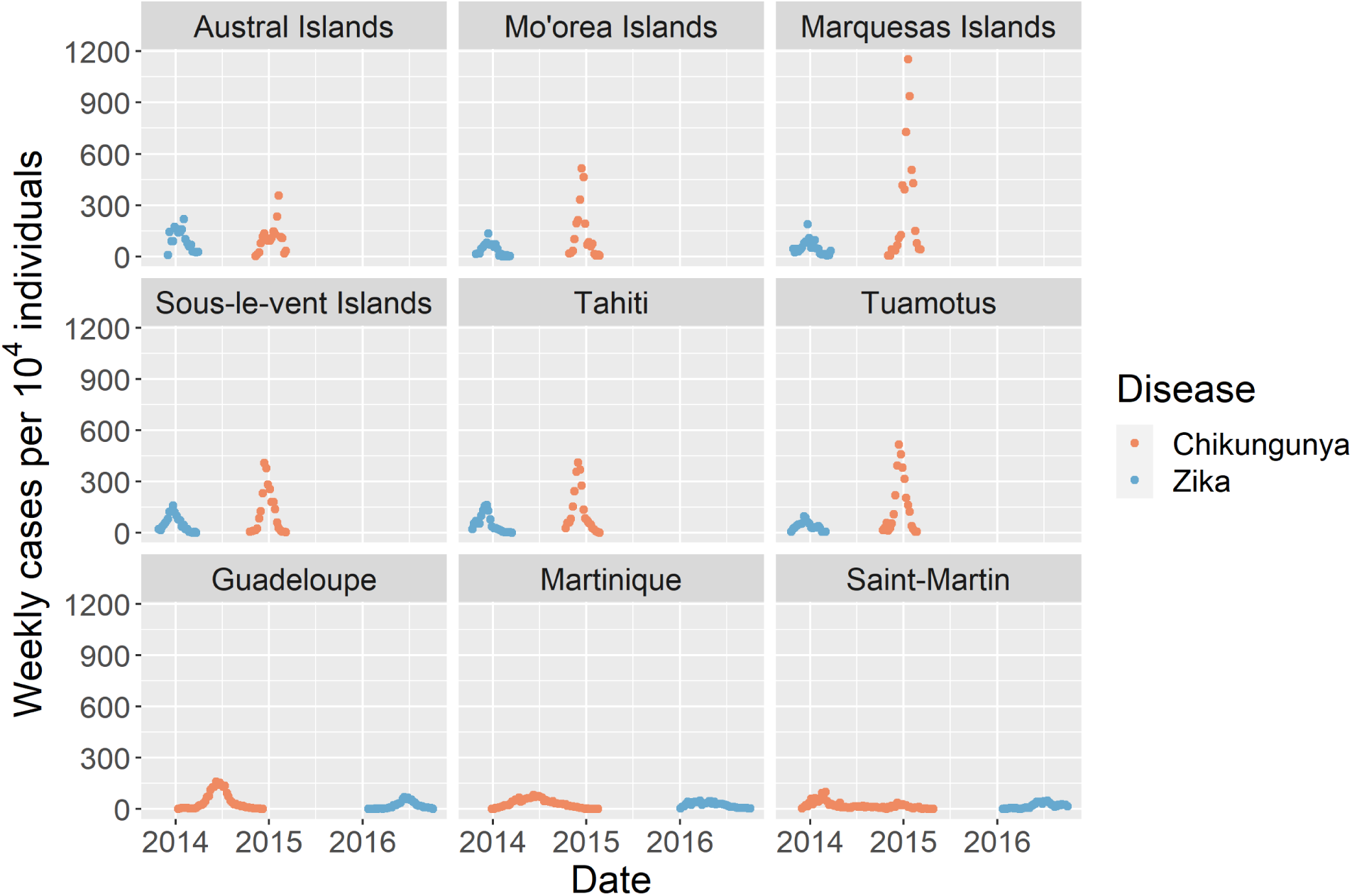
Normalized incidence of Chikungunya and Zika fevers in nine different regions during the outbreaks. The outbreak incidence data for Chikungunya and Zika are shown by orange and blue points, respectively.

## 2 Method

In order to explore the necessity of incorporating both mosquito population dynamics and local temperature data in the models for arbovirus outbreaks, we developed four deterministic models to describe the mean number of new cases in each week. We then compared these four models based on their goodness of fit to recorded case data. After identifying our “best model,” we used it to estimate biologically meaningful parameters, in order to understand the transmissions of Chikungunya and Zika fevers. We used a maximum likelihood approach, implemented in R, followed by Akaike Information Criterion analysis for model comparison and selection. Finally, we used Bayesian inference to obtain the posterior values of the parameters and posterior predictions of the current data.

### 2.1 Data description

The incidence data consist of groups of successive weekly cases of Chikungunya and Zika in nine different regions, including six islands and archipelagoes of French Polynesia (Austral, Marquesas, Mo’orea, Sous-le-vent, Tahiti, and Tuamotus) and three regions in the French West Indies (Guadeloupe, Martinique, and Saint-Martin) [3]. The recorded dates range from October 13, 2013 to October 2, 2016, and the weekly data for both Chikungunya epidemics and Zika epidemics are available for each region, as shown in Figure 1. The weekly average temperature in celsius is also available for every region and outbreak period, as shown in Appendix Figure S1.

### 2.2 Model description

We described the number of weekly new cases using a Poisson distribution. Since the number of new cases is much smaller than the total population size, we assumed that the propensity of the disease transmission reaction does not change too much in a week. Thus we employed the so-called tau-leaping approximation for the underlying continuous-time, discrete-state Markov process [25, 26]. We assume the number of new cases in week *i*, Δ*I_i_,* follows a Poisson distribution with mean *λ_i_ × t*. We adopt one week as our basic time unit, thus *t* equals 1. Consequently,

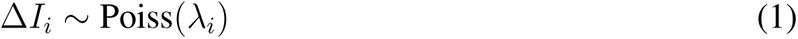

We consider four different models for determining the parameter *λ_i_*, developed in the following subsections: mass-action model, mosquito model, temperature model, and mosquito-temperature model.

#### 2.2.1 Mass-action model

Following [27], we model the transmission of a disease in a homogeneously mixed population using the law of mass action, with the number of new infections being proportional to the fraction of the remaining susceptible individuals. In our simplest model, we describe the mean number of new cases per week as follows:

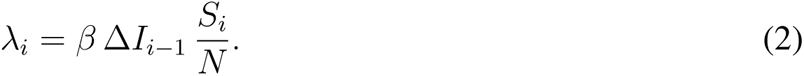

Here *S_i_* represents the number of susceptible individuals at the start of the current week (*i*) and Δ*I_i−_*_1_ represents and the number of newly infected individuals in the previous week (*i −* 1). The initial population size, *N*, is treated as a constant. The parameter *β* represents the average number of transmissions per week per infected individual if the entire population were susceptible. We assume that the new infections in week (*i*) are caused by the newly infected individuals in week (*i −* 1). That is, we assume that the infected individuals generated from the weeks before the previous week have been removed from the general population, either through recovery, or through admittance to hospitals. These individuals comprise a removed class that we take to be an absorbing state. For this model, the basic reproduction number *R*_0_ of the disease, i.e. the average number of secondary transmissions from one infected person in a total susceptible population is expressed as:

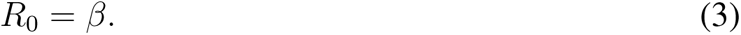

#### 2.2.2 Mosquito model

Both the Chikungunya virus and the Zika virus are transmitted via mosquito. To explore the effect of mosquito population dynamic on Chikungunya and Zika transmission, we augmented the preceeding model so as to consider the dynamics of the infected mosquitoes and susceptible mosquitoes in each week. For simplicity, in this model we assume that the total population of the mosquito remains constant during an outbreak in a specific region. The “Mosquito model” variables are the number of susceptible mosquitoes *M_s_* and the number of infected/virus-carrying mosquitoes *M_i_*. By assumption, *M_s_*+ *M_i_* = *N_M_*, where *N_M_* represents the total mosquito population size. The incidence of mosquito biting is assumed to follow the law of mass action. We define *β* to be the average number of mosquitoes that bite on each infected person per day if all the mosquitoes are susceptible. In addition, the infected mosquitoes can become virus-free again with per capita rate *α*. We write the kinetics of the mosquito population as:

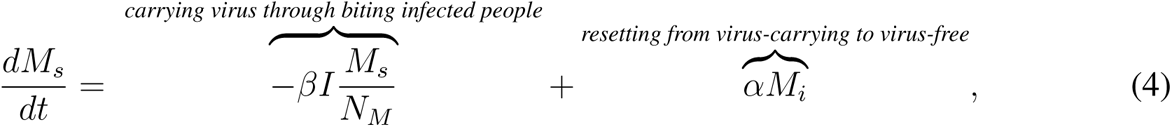

Since we consider the total mosquito population size to be constant, *M_i_* obeys a scalar differential equation:

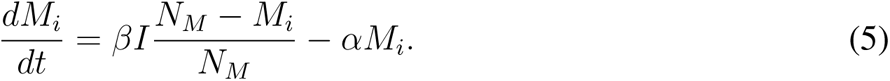

We assume that the infected mosquito population size converges quickly enough to its equilibrium each week that we can replace the infected population with its steady-state value:

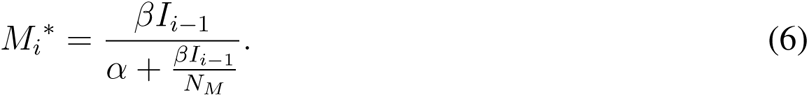

Therefore we extend the previous mass-action model (2) so that the average number of new cases in week *i* can be described as

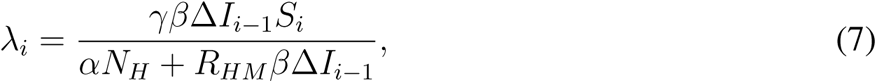

where *N_H_* is the initial human population size, and *R_HM_* = *N_H_/N_M_* represents the initial human-to-mosquito ratio. Moreover, *γ* represents the average number of transmissions per week per infected mosquito.

To derive the *R*_0_ of the disease for this model, we first calculated the average number of infected mosquitoes produced by one infected person in the next week under the assumption that the human population size is sufficiently large as below:

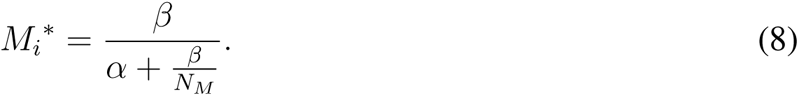

Then, the expression for *R*_0_, can be obtained as:

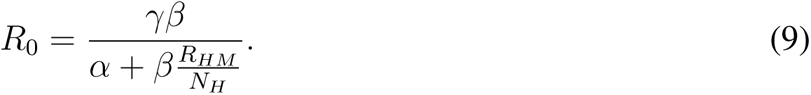

#### 2.2.3 Temperature model

To study the effects of temperature on the transmissions of Chikungunya and Zika fevers, we extended the mass-action model to incorporate the weekly average temperature *wt*. To the best of our knowledge, the effect of temperature on the transmission rate *β* has not been reported. For the same of simplicity, we adopt a linear Ansatz to account for the positive associations of the temperature with the host-seeking ability, biting rate, and development rate of the mosquitoes [28].

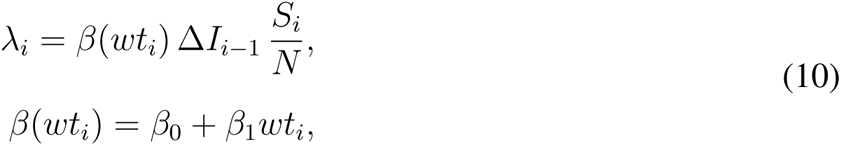

where the transmission rate *β* in week *i* is a linear function of the average weekly temperature in week *i* (*i ≥* 0), *wt_i_*. Under this assumption, the *R*_0_ of the disease is defined as

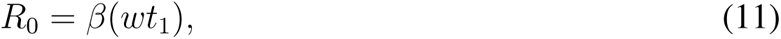

where *wt*_1_ is the average temperature in the second week of the outbreak.

#### 2.2.4 Mosquito-temperature model

Previous studies show that temperature can affect the mosquito survival and activity. In particular, the immature stages of mosquito (e.g. larvae and pupae) has the optimal survival at 25-30 ℃, and the mortality rates will dramatically increase at extreme temperatures [29, 30]. Temperature can also affect the mean duration of the extrinsic incubation period (EIP), and Johansson et al. models the EIP in the form *a × e^b×^*^(^*^T−^*^28)^ [31]. Temperature can also affect the mean duration of the gonotrophic cycle (GC), and the relationship can be described by the following quadratic equation *GC* = 56.64 *−* 3.736 *∗ T* + 0.064 *∗ T* ^2^ [3, 32]. Since GC includes the mosquito biting, one can expect that a decrease in the average duration of GC will lead to an increase in the mosquito attack rate. Thus, we assumed that the mosquito attack rate is proportional to the reciprocal of the mean duration of GC, and to incorporate the effect of temperature on the mosquito biting, we set both parameter *β* and *γ* to be dependent on the mean weekly temperature *wt_i_*.

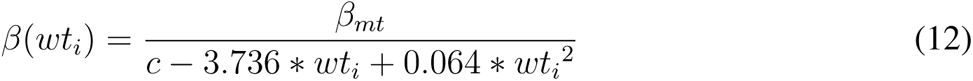

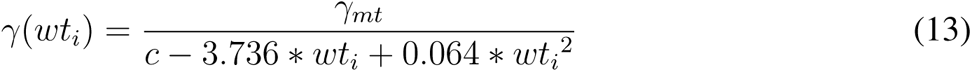

With the framework as the mosquito model, the mosquito temperature model has all the *β* and *γ* replaced by the *β*(*wt_i_*) and *γ*(*wt_i_*), which results in the formula below:

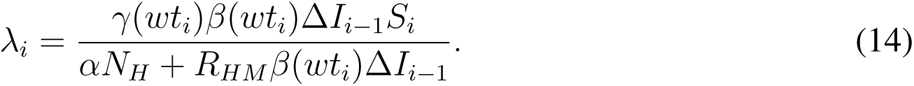

The equation for *R*_0_ has the similar form as equation 9, by replacing all the *β* and *γ* with *β*(*wt*_1_) and *γ*(*wt*_1_), where *wt*_1_ is the average temperature in the second week of the outbreak.

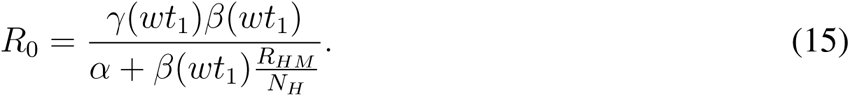

### 2.3 Model comparison, parameter estimation and simulation

The number of contacts between people and mosquitoes is likely to be large in French Polynesia. At the same time, the probability of successful disease transmission *per contact event* should be small (due to various factors such as the mosquito’s incubation period and the host’s immunity). Consequently, the Poisson distribution appears to be a reasonable (and parsimonious) choice for modeling the weekly incidence rates. For the model comparison and initial model simulations, therefore, we used the Poisson distribution for weekly cases and the maximum likelihood approach to derive the loglikelihoods of the different models across different groups of data.

Since the Poisson distribution is not overdispersed, we compared models based on on their abilities to describe the variation by using their core parameters. In other words, by assuming that the process is less stochastic, we can fully compare the predictive powers of the different models. Using overdispersion in model comparison might cause a low-bias model to be less distinguishable from a high-bias model because the mean has less influence on the likelihood than the variance.

We used the Akaike Information Criterion (AIC) [33, 34] to evaluate and compare the models. By convention, the AIC is calculated as *AIC* = *−*2*L* + 2*k,* where L and k are the log-likelihood and number of parameters, respectively. The AIC metric is an indirect estimation of the test error of a model, and it penalizes the model for having more parameters due to a higher chance of overfitting. Due to the limited amount of data available, performing validation or cross-validation may overestimate the test error. Also, since our models are not purposed primarily for accurate prediction, we consider AIC to be sufficient for model selection.

We found that model simulations using the Poisson distribution tend to underestimate the variance of the field data. As shown in Appendix B (Figure S2), the data is more dispersed than any of the four models would predict. Therefore, it appears that the Poisson distribution fails to capture all sources of stochasticity in the disease transmission process. In order to construct more realistic simulations, and to pursue more accurate parameter estimation, we used replaced the Poisson distribution with a Normal distribution (with some overdispersion) in order to approximate the number of weekly cases. The additional stochasticity could arise from unspecified human, mosquito, and (or) human-mosquito processes. For instance, the human recovery period and the weekly infected mosquito population are both random quantities, but are treated as deterministic in the Poisson model.

Thus, we introduce a scaling factor *σ* to account for the various sources of process variations that are ignored in the Poisson model, as well as (random) errors due to the *τ* -leaping assumption that the propensities of the reactions stay unchanged in one week. We write the incidence of new cases as:

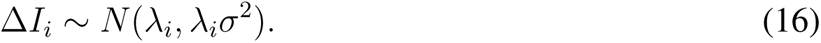

To ensure that our model comparison result and best-model choice still hold after replacing the Poisson distribution with the Gaussian distribution, we repeated our model comparison for all four models using the Gaussian likelihood function across different values of *σ*. As shown in Appendix C (Figure S4), our best-model choice holds for all *σ* less than 10. For larger values of the disperson parameter *σ*, the four models become indistinguishable from each other. However, for every region except Tahiti, the estimated posterior distributions of *σ* for both diseases indicate that *σ* is likely to be below 10. The larger *σ* value obtained from the Tahiti data possibly suggests that some other covariates at this particular region contribute more variability than the mosquito-human transmission process. Thus, the conversion from the Poisson distribution to the Gaussian distribution does not typically alter the conclusions from the previous model comparison results with *σ* = 1.

#### Computational methods

For this inferential study, we used *Stan*, a programming language for MCMC-based Bayesian inference, to obtain the posterior distributions and 90% credible intervals of the parameters (https://mc-stan.org). The posterior predictive distributions are then obtained to check the agreement of the model prediction and the training data.

## 3 Result

### 3.1 Comparing models

In order to select the model that best balances parsimony with accurate data fitting, we calibrated all four models with respect to the data from all nine regions. We then applied the AIC test for model comparison and model selection. Table 1 shows the results of the AIC tests evaluated for outbreak data from different regions. We compute the ΔAIC of each model by subtracting the AIC of the best model from the AIC of the current model. A smaller ΔAIC corresponds to shorter distance to the best model, and the model with zero ΔAIC is the best model. As shown in Table 1, incorporating mosquito dynamics into the model results in a significant improvement in the accuracy of prediction, which is supported by both diseases across all the regions. In contrast, the mosquito-temperature model is only justified by both diseases in the Austral Islands and the Marquesas Islands, suggesting that incorporating the effect of temperature on mosquito biting gives only a limited enhancement of the predictive power. Figure S2 (Appendix B) compares simulations of the four calibrated models to Austral Chikungunya incidence data. Since the mosquito model is more strongly justified than the mass-action and temperature model, and has similar predictive power but fewer parameters and predictive variables than the mosquito-temperature model, we conclude that the mosquito model is the best model among all four models in this study.

**Table 1:**
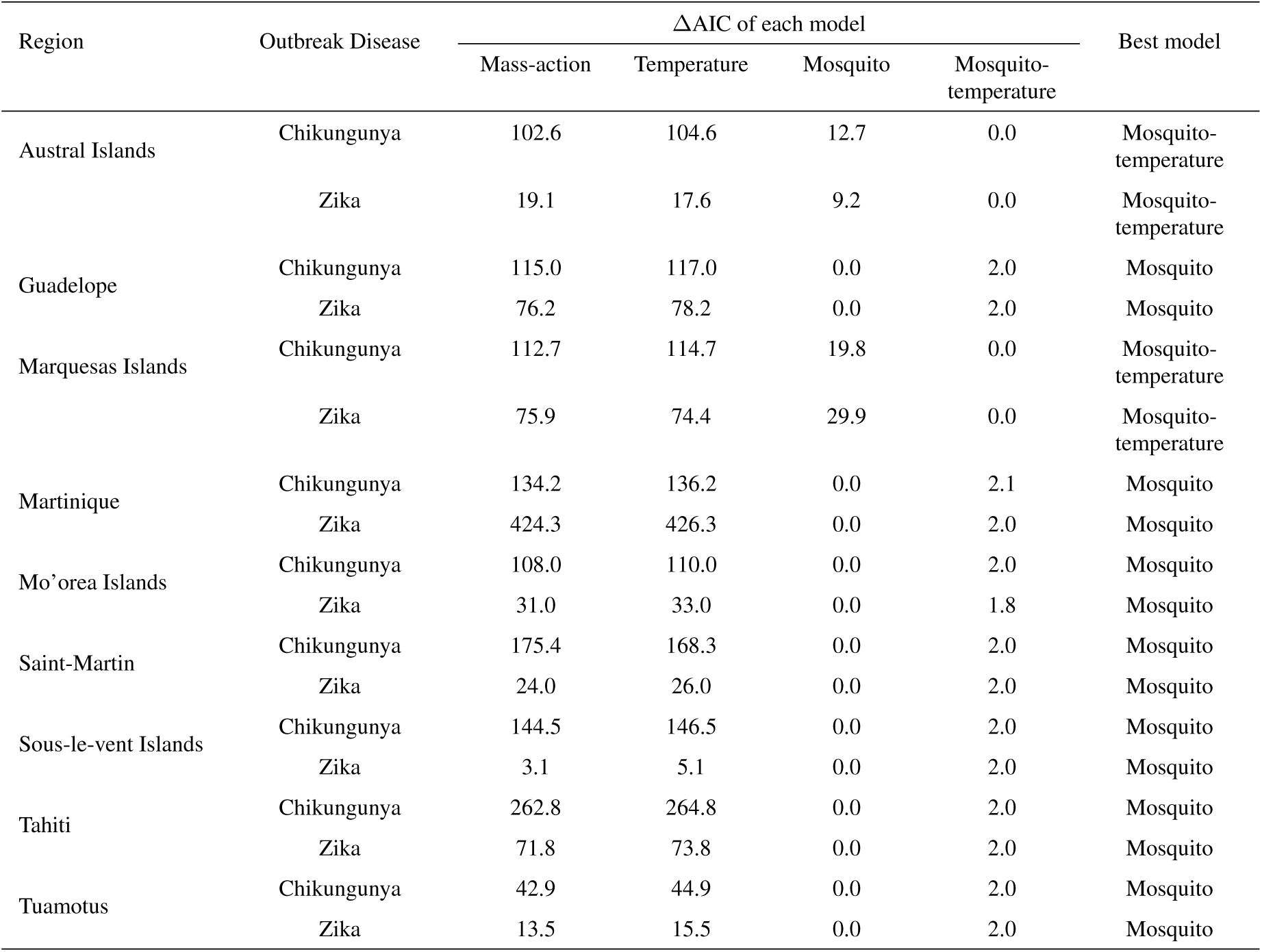
Model comparison summary: AIC test comparison the four models

**Table 2:** Median and 90% credible intervals of the parameters in the mosquito model

| Region | Virus | Parameter | | | | $R_0$ |
| --- | --- | --- | --- | --- | --- | --- |
| | | $\alpha(\text{time}^{-1})$ | $\beta(\text{time}^{-1})$ | $R_{HM}$ | $\gamma(\text{week}^{-1})$ | |
| Austral Islands | Chikungunya | 2.86 [0.61; 6.62] | 2.18 [0.45; 6.40] | 3.47 [0.51; 8.03] | 1.63 [0.34; 5.34] | 1.20 [0.90; 1.57] |
|  | Zika | 3.07 [0.71; 6.83] | 1.97 [0.43; 5.96] | 3.10 [0.30; 7.60] | 1.67 [0.36; 5.41] | 1.07 [0.89; 1.29] |
| Guadelope | Chikungunya | 3.12 [0.82; 6.66] | 1.97 [0.44; 5.66] | 3.19 [0.42; 7.61] | 1.79 [0.51; 5.50] | 1.15 [1.08; 1.23] |
|  | Zika | 3.06 [0.69; 6.71] | 1.97 [0.39; 6.04] | 3.11 [0.38; 7.44] | 1.66 [0.35; 5.66] | 1.06 [0.97; 1.15] |
| Marquesas Islands | Chikungunya | 3.03 [0.84; 6.68] | 2.31 [0.53; 6.04] | 3.33 [0.59; 7.40] | 2.28 [0.77; 5.65] | 1.67 [1.31; 2.21] |
|  | Zika | 3.00 [0.62; 6.67] | 2.02 [0.44; 6.15] | 3.32 [0.47; 7.70] | 1.63 [0.29; 5.60] | 1.09 [0.82; 1.37] |
| Martinique | Chikungunya | 2.76 [0.58; 6.51] | 2.27 [0.47; 6.61] | 3.57 [0.53; 7.88] | 1.42 [0.27; 5.17] | 1.13 [1.07; 1.23] |
|  | Zika | 2.84 [0.49; 6.86] | 2.15 [0.39; 6.29] | 3.34 [0.48; 7.68] | 1.56 [0.21; 5.33] | 1.06 [0.98; 1.18] |
| Mo'orea Islands | Chikungunya | 3.06 [0.82; 6.53] | 2.08 [0.45; 6.06] | 3.33 [0.52; 7.53] | 1.87 [0.51; 5.56] | 1.25 [0.96; 1.61] |
|  | Zika | 2.97 [0.68; 6.93] | 1.92 [0.41; 5.97] | 3.07 [0.33; 7.75] | 1.69 [0.30; 5.63] | 1.04 [0.80; 1.31] |
| Saint-Martin | Chikungunya | 2.84 [0.49; 6.49] | 2.19 [0.48; 6.50] | 3.44 [0.45; 7.89] | 1.51 [0.19; 5.45] | 1.11 [0.98; 1.26] |
|  | Zika | 2.86 [0.50; 6.55] | 2.09 [0.43; 6.27] | 3.22 [0.43; 7.69] | 1.58 [0.20; 5.55] | 1.07 [0.97; 1.20] |
| Sous-le-vent Islands | Chikungunya | 2.88 [0.67; 6.45] | 2.24 [0.49; 6.44] | 3.53 [0.51; 7.72] | 1.57 [0.42; 5.45] | 1.25 [1.03; 1.61] |
|  | Zika | 3.11 [0.77; 6.95] | 2.02 [0.39; 5.91] | 3.08 [0.33; 7.52] | 1.74 [0.43; 5.48] | 1.07 [0.93; 1.23] |
| Tahiti | Chikungunya | 3.06 [0.73; 6.85] | 1.99 [0.41; 5.92] | 3.05 [0.34; 7.45] | 1.86 [0.56; 5.72] | 1.22 [1.05; 1.44] |
|  | Zika | 3.10 [0.75; 6.91] | 1.86 [0.40; 5.91] | 3.16 [0.31; 7.44] | 1.81 [0.41; 5.76] | 1.08 [0.93; 1.24] |
| Tuamotus | Chikungunya | 2.87 [0.65; 6.52] | 2.14 [0.43; 5.97] | 3.30 [0.49; 7.70] | 1.80 [0.54; 5.75] | 1.32 [1.10; 1.65] |
|  | Zika | 3.06 [0.58; 6.80] | 1.97 [0.40; 6.13] | 3.21 [0.50; 7.55] | 1.72 [0.28; 5.46] | 1.06 [0.88; 1.28] |

### 3.2 Biological insight about Chikungunya and Zika based on parameter estimation

#### 3.2.1 Basic reproductive number *R***_0_**

The goal of this study is to compare the scales of Zika and Chikungunya outbreaks among all nine regions, and to investigate differences in the transmission dynamics of these two diseases. Our results from the best-fit model (Figure 2) show that (i) the Zika *R*_0_ is similar in each region; (ii) Chikungunya possesses a higher *R*_0_ value than Zika universally in all regions; (iii) Marquesas Islands possesses significantly higher Chikungunya *R*_0_ value than all other regions. These observations are consistent with the outbreak data shown in Figure 1. Thus they serve to support our model: the Zika outbreak in each region is of a similar scale and smaller than the Chikungunya outbreak, and the Marquesas Islands experienced the largest Chikungunya outbreak among all nine regions.

**Figure 2:**
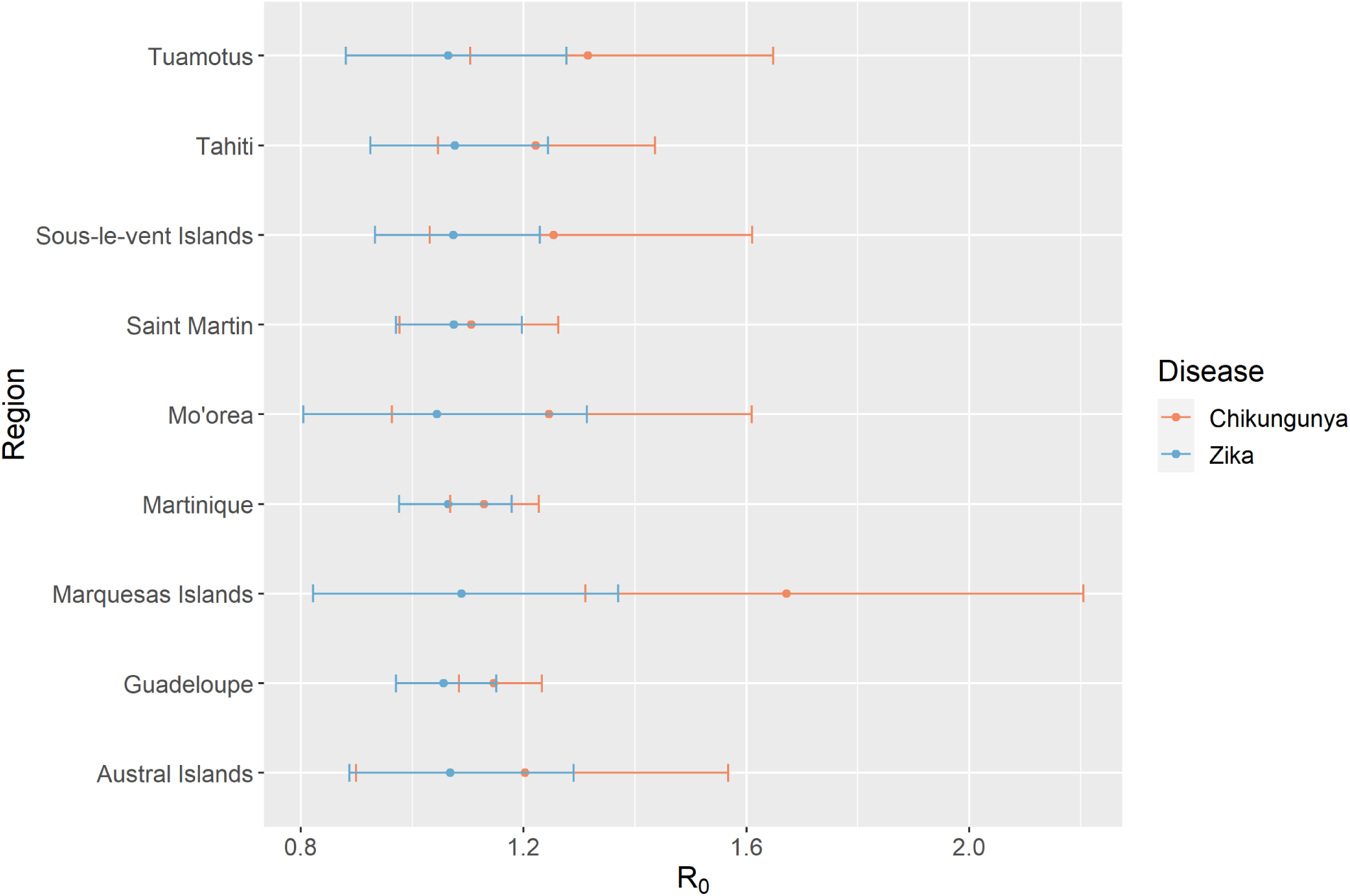
Basic reproductive number *R*_0_ estimated from the mosquito model (section 2.2.2). Each dot refers to the median value of estimated *R*_0_ and the whiskers represent the 90% credible interval of the estimation.

In addition, to better understand the effects of the parameters of the mosquito model on the *R*_0_, we evaluated the elasticities for all the parameters at the fitted parameter values for the Chikun-gunya outbreak in the mosquito model in the Austral islands, shown in Figure S3 (Appendix B).

#### 3.2.2 Transmission parameters *β, γ, R_HM_* and *α*

*Aedes aegypti* and *Aedes albopictus* are the two primary vectors responsible for the transmission of both viruses. Though sharing a few common characteristics, the two mosquito species should contribute differently to the transmission of each disease, as they still differ in the feeding preference, thermal-dependent life cycle, extrinsic incubation period, and transmission ability, inter alia. Further, the prevalence of the two mosquito species could differ significantly among regions, resulting in different outbreak potentials of vector-borne diseases. Due to the lack of species-specific transmission parameter estimation, our model accounts for the overall impact of *Aedes* mosquitoes. Therefore, the model parameters estimated from each fitting only represent a combined effect of both species and thus should be different among regions.

##### Chikungunya and Zika outbreak sizes in the same region

Although all nine regions experienced various scales of Zika and Chikungunya outbreaks, our parameter estimation (Figure 3) indicates that the specific reason for the unequal outbreak sizes could be distinctive from region to region. Marquesas Islands is estimated to possess human-to-mosquito ratios (*R_HM_*) and the infectious mosquito turnover rates (*α*) that are close for Zika and Chikungunya, meaning that the components of mosquito species for the transmission of both viruses are similar. At the same time, the Chikungunya outbreak in this region was significantly larger. Therefore, the larger outbreak is attributable to other parameters, namely the transmission rates between mosquitoes and humans (*β, γ*). This analysis suggests a high biting rate and poor mosquito bite prevention in the Marquesas Islands. In contrast, the larger Chikungunya outbreak in Sous-le-vent Islands appears to have been caused by a slower infectious mosquito turnover rate and higher human-to-mosquito transmission rate, even though more mosquitoes spread Zika than Chikungunya.

**Figure 3:**
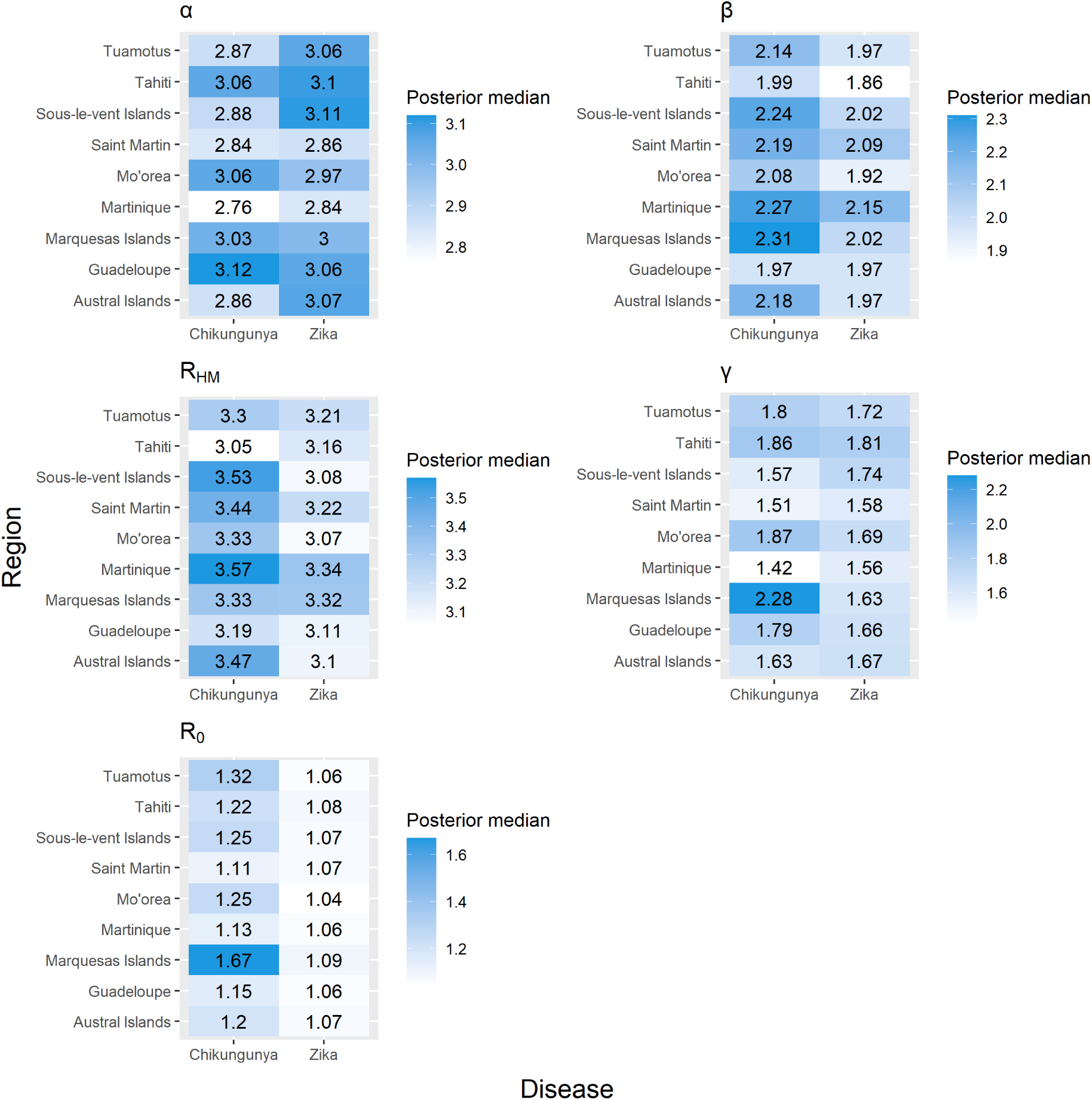
Parameter estimation of mosquito model (section 2.2.2) for both diseases at nine regions in French Polynesia.

##### Chikungunya and Zika outbreak sizes among regions

The outbreak sizes and *R*_0_ of the Zika virus vary minimally among all regions, and so do the estimated model parameters. The homogeneous estimations on *β* and *γ* suggest that the effective biting rates of mosquitoes transmitting the Zika virus are similar among most regions. Further, the estimations on the human-to-mosquito ratio for Zika transmission are close in many regions, which indicates the component of mosquitoes that transmit Zika is similar among these regions. These findings suggest that a uniform vector control strategy could be applied universally to all regions to prevent future Zika virus outbreaks.

On the other hand, the parameter estimations are diverse for Chikungunya in most regions. For example, the Marquesas Islands possess the highest values of *β* and *γ* but a moderate value of *R_HM_*, which indicates the largest Chikungunya outbreak is attributable to a more prominent component of mosquito species that transmits Chikungunya more effectively, not to a small human-to-mosquito ratio. Therefore, in order to manage future Chikungunya outbreaks, one could employ vector control plans that target the mosquito species with the most efficient transmission ability of the virus.

### 3.3 Model simulations and comparison with field data

Figures 4 and 5 illustrate the simulated average weekly cases versus time for Chikungunya and Zika fevers, respectively. The green shaded area gives the 90% credible interval for the number of simulated cases per week, all of which matched the reported incidence data very well. Hence, the calibrated “best model” - the mosquito model - captures most features of the outbreaks.

**Figure 4:**
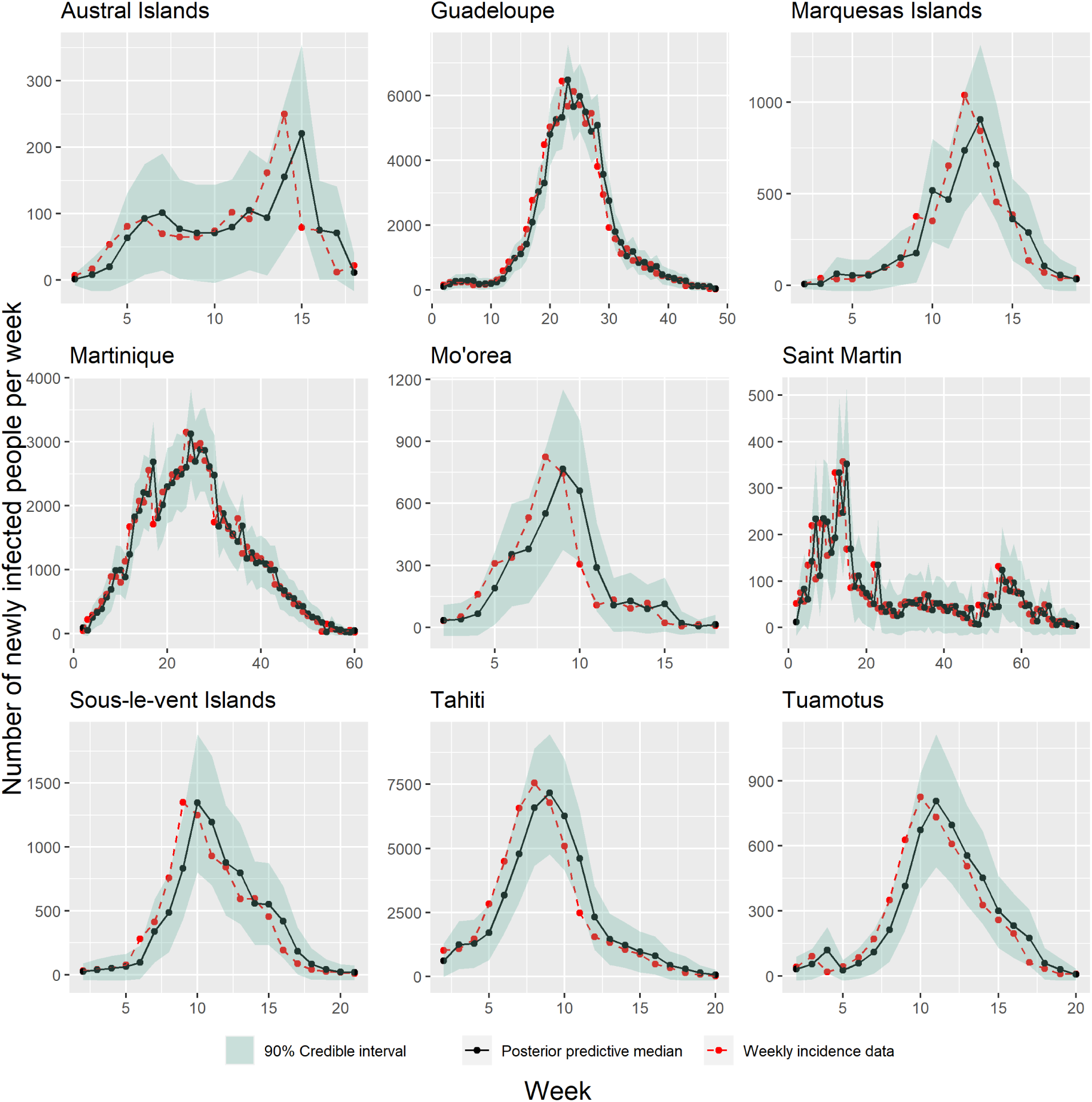
Mosquito model (section 2.2.2) fittings with Gaussian likelihood function to the Chikungunya disease incidence data in all nine regions.

**Figure 5:**
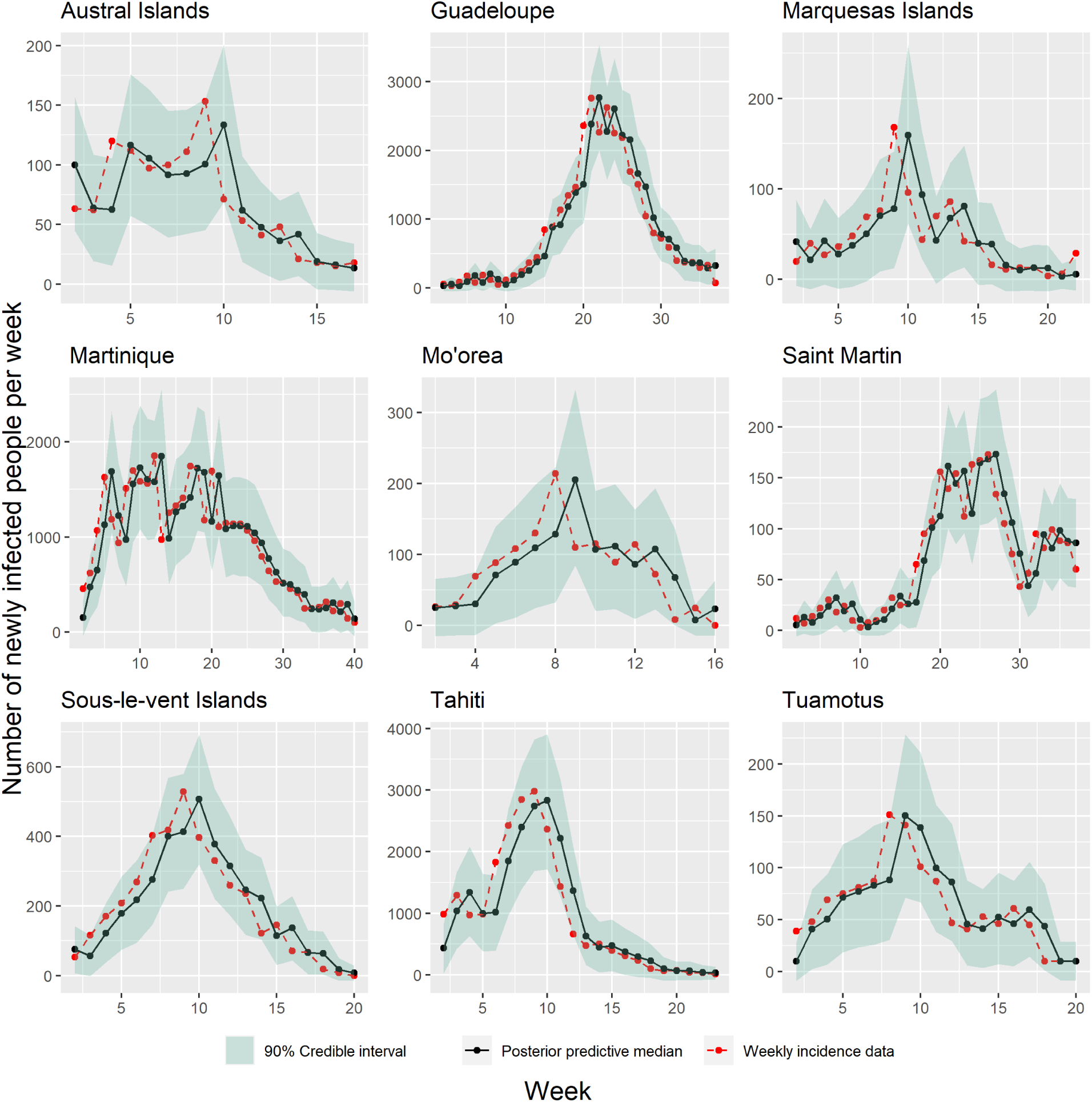
Mosquito model (section 2.2.2) fittings with Gaussian likelihood function to the Zika disease incidence data in all nine regions

## 4 Discussion

The Chikungunya and Zika diseases have threatened public health in many regions, and the rapid expansions of both diseases could potentially lead to future global pandemics [35]. In addition to efforts to develop drugs and vaccines, mathematical modeling is an important tool for disease-control planning. The usefulness of modeling depends on its ability to predict the propagation of infectious diseases and identify factors that can affect patterns of spread [36, 37]. Ideally, an inferential model should capture the key factors influencing outbreaks, but contain as few parameters as possible in order to save computational power and ensure robust parameter estimation.

Studies have shown that French Polynesia is at high risk for having Chikungunya and Zika outbreaks due to the presence of two local mosquito species: *Aedes aegypti* and *Aedes polynesiensis* [38, 39]. A study has also shown that though sharing a few common characteristics, these two species differ in feeding habits, thermal-dependent life cycle, extrinsic incubation period, and transmission ability [2]. Moreover, they are highly competent in transmitting the arboviral diseases. Hence, it is likely that these two species contribute differently to the transmission of each disease. In addition, the hot and rainy season in French Polynesia that continues from October to March favors the reproduction of the mosquitoes [40]. Hence, to better understand the commonalities and differences among these two *Aedes*-transmitted diseases, we conducted a joint analysis of Chikungunya and Zika outbreaks in French Polynesia, building on common aspects in location and vectorial transmission.

Firstly, to investigate the necessity of including mosquito population dynamics and the local temperature data in models of the arboviral outbreaks, we developed four deterministic models with and without the above two features as follows: a mass-action model, a mosquito model, a temperature model, and a mosquito-temperature model. We assumed the Poisson distribution for the weekly cases. One advantage of such choice is that it could make the low-bias models more distinguishable from the high-bias models than overdispersed distributions. We calibrated all four models using weekly incidence data of Chikungunya and Zika in nine different regions in French Polynesia, and we applied the AIC test for model comparison and model selection. Our results (Table 1) suggested that mosquitoes play a crucial role in the spread of arboviral diseases. While the effect of temperature on the spread of arboviral diseases varies across different regions, the temperature does not appear to play as significant a role as the mosquito dynamics. In particular, the effect of temperature on mosquito biting is one of the dominant factors that determines the rate of disease spread in the Austral islands and Marquesas islands, which is not true in the other regions. This difference probably arises because the temperature also affects aspects other than the mosquito biting, such as the mosquito population and extrinsic incubation period (EIP) [41, 42]. In the present analysis, we concluded that incorporating the mosquito population dynamics in the transmission of arbovirus modeling is essential to enhance the predictive power, while the temperature impact may not be necessary for understanding past outbreaks in French Polynesia.

Our study included some but not all factors that could influence patterns of disease spread. Although we have parameterized some of the key factors involved in Arboviral disease transmission, we omitted other mosquito factors, such as the EIP and gonotrophic cycle, as well as human factors such as human behavioral changes through the progression of the disease outbreak, asymptomatic cases that can cause additional disease transmissions, the incubation period and immunity. Although these factors were excluded from the present study, they could be considered for more accurate model prediction in future studies. Here, we used the mosquito model as our best model among the four models considered. Overall, our calibrated “best model” captured most of the features of the outbreaks (Figures 4 and 5).

As we know, the basic reproduction number, *R*_0_, defined as the average number of secondary transmissions from one infected person in an otherwise susceptible population, has important implications for mitigation efforts needed to bring an epidemic under control. We observed that (shown in Figure 2): (i) the Zika *R*_0_ is similar in each region; (ii) Chikungunya possesses a higher *R*_0_ value than Zika universally in all regions; (iii) Marquesas Islands possesses significantly higher Chikungunya *R*_0_ value than all other regions. These observations are consistent with the outbreak data shown in Figure 1. We further analyzed the following four parameter values to obtain biological insight of Chikungunya and Zika diseases: mosquito biting rate on infected people (*β*), disease transmission rate from infected mosquitoes to susceptible people (*γ*), human-to-mosquito ratio (*R_HM_*), and the infectious mosquito turnover rate (*α*). It is worth noting that the parameters *β* and *γ* represent the joint effects of many factors. Parameter *β* represents the combined effect of the mosquito biting and the incubation period on the rate at which the susceptible mosquito becomes infected. Similarly, parameter *γ* represents the combined effect of the mosquito biting and the transmission probability on the rate of disease transmission from the mosquito to human. Our results (Figure 3) indicate that the specific reason for the unequal outbreak sizes of Zika and Chikungunya could vary from region to region. For example, the large Chikungunya outbreak in the Marquesas Islands is probably attributable to the transmission rates between mosquitoes and humans (*β, γ*), which indicates a high biting rate and poor mosquito bite prevention in the Marquesas Islands. In contrast, the larger Chikungunya outbreak in Sous-le-vent Islands is caused by a slower infectious mosquito turnover rate and higher human-to-mosquito transmission rate, even though more mosquitoes spread Zika than Chikungunya. Moreover, the parameter estimations are diverse for Chikungunya in most regions. On the other hand, the outbreak sizes and *R*_0_ of the Zika virus vary minimally among all regions, and so do the estimated model parameters. These findings suggest that universal vector control plans will help prevent future Zika outbreaks, but targeted control plans focusing on specific mosquito species could aid the prevention of Chikungunya outbreaks.

Future investigations could explore how population fluctuations and behavioral changes of the mosquito population are influenced by seasonal factors. In addition, a better understanding of how weather affects the mosquitoes during the disease transmission could further strengthen disease-control efforts.

## Data Availability

All data produced in the present study are available upon reasonable request to the authors.

## Acknowledgments

This work was supported in part by National Science Foundation grant DMS-2052109 to PJT, grants DMS-1853622 and DMS-2052648 to XH. PJT acknowledges research support from the Oberlin College Department of Mathematics. QH acknowledges research support from the College of Wooster.

## Appendix

## Appendix A: Temperature data in nine different regions

**Figure S1:**
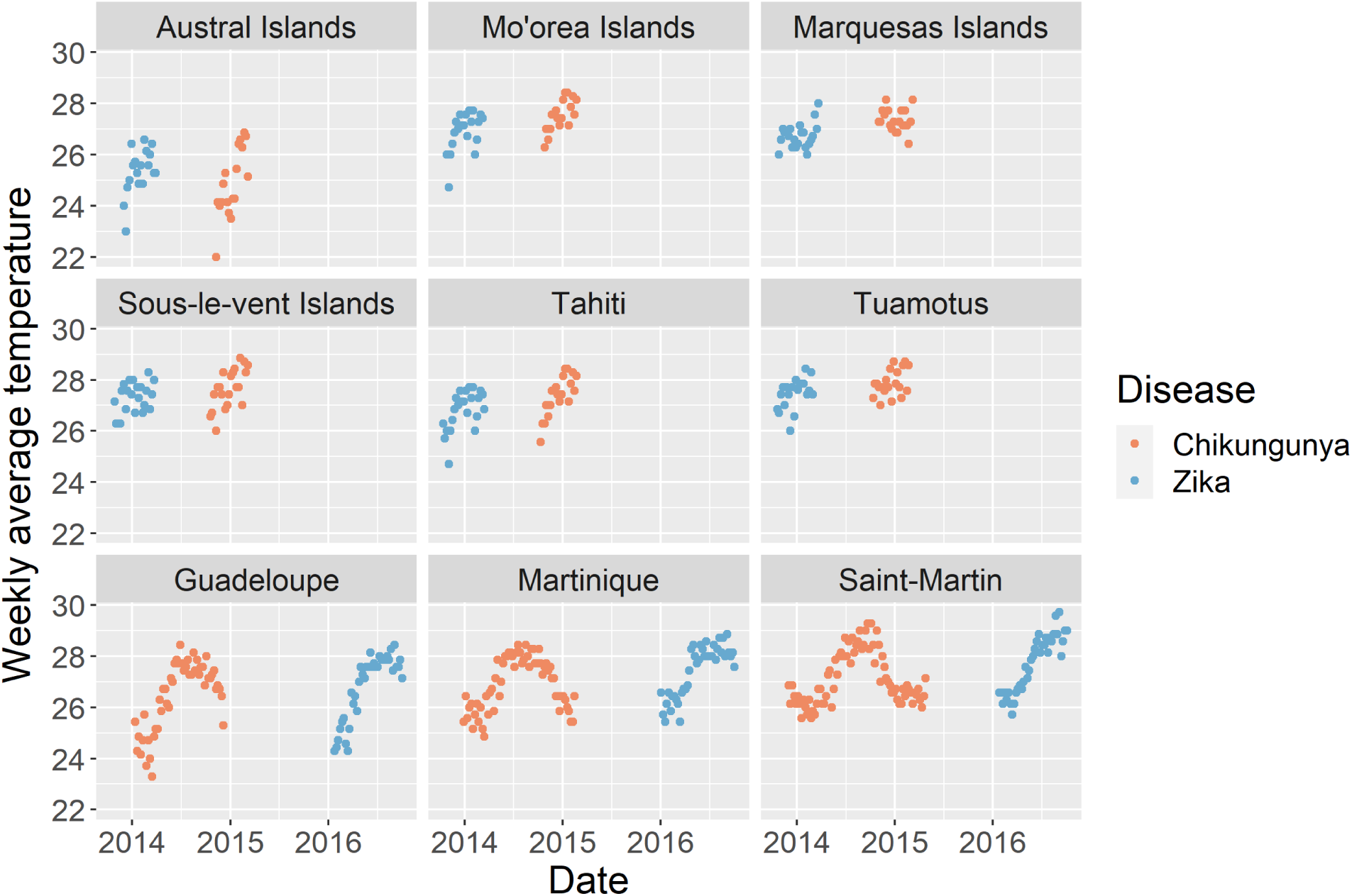
Weekly average temperature data during the outbreaks of both diseases at nine regions in French Polynesia. The outbreak temperature data for Chikungunya and Zika are shown by orange and blue points, respectively. The temperature data is used as an input variable in the temperature (section 2.2.3) and mosquito-temperature models (section 2.2.4).

## Appendix B: Model comparison

### Simulations of the four calibrated models and comparison to Austral Chikungunya incidence data

As shown in Figure S2, the upper and lower limits of the 1000 randomly simulated samples bound most of the observed data points, indicating that all four models can capture most of the features of the Chikungunya outbreak in the Austral islands. Specifically, the mass-action model and temperature model produce similar predictions on the weekly mean cases (Figure S2A and B). The mosquito model and the mosquito-temperature model generate much different predictions than the other two models (Figure S2C and D), and their predicted weekly means are much closer to the real data compared to the mass-action model and temperature model. The similar predictions between mosquito and mosquito-temperature model suggest that incorporating temperature does not qualitatively change predictive power. In addition, from week 1 to week 5, the predictions of the mosquito model and mosquito-temperature model fit the data better than the those of the other two models, indicating the importance of mosquito in the early stage of the disease spread. For the portion from week 6 to week 11, the mosquito-temperature model’s prediction fits the data better than that of the mosquito model, suggesting that the effect of temperature on mosquito biting can affect the rate of disease spread. All four models fail to accurately predict the peak between week 12 and week 14, indicating additional stochasticity involved in the disease transmission process that has not been formulated into the models.

**Figure S2:**
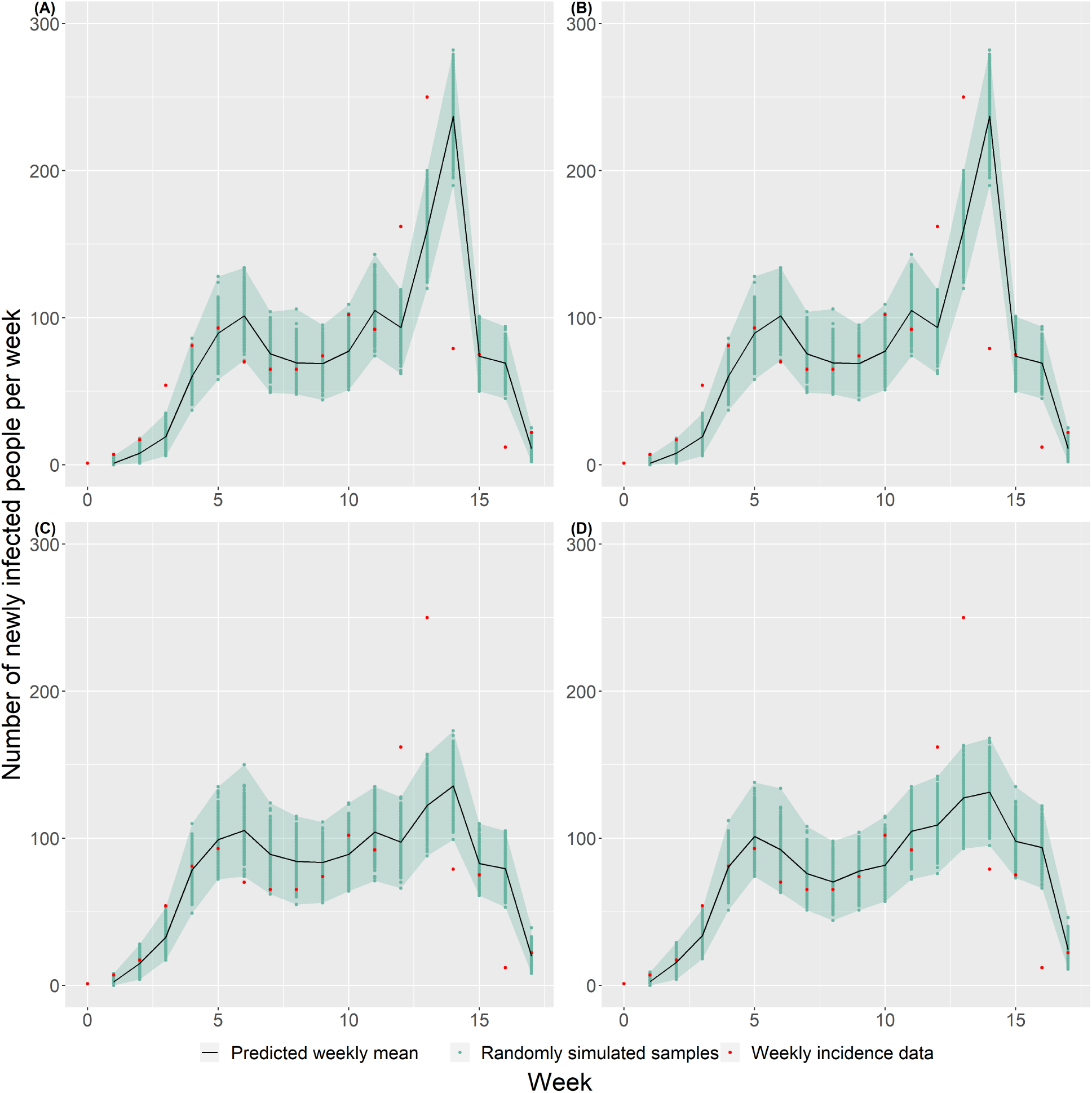
Simulations of the four calibrated models and Austral Chikungunya incidence data: (A) Mass-action model, (B) Temperature model, (C) Mosquito model, (D) Mosquito-temperature model. 1000 random simulations of each model are shown by the green dots in the corresponding sub-figure. The margin of the green-shaded region in each sub-figure indicates the upper and lower bounds of these 1000 simulations. Each model predicts the expected number of weekly cases for each week, which is shown by the black curve in each sub-figure.

### Local elasticity analysis

To better understand the effects of the parameters of the mosquito model on the *R*_0_, we calculated the local elasticity for each parameter by using this equation:

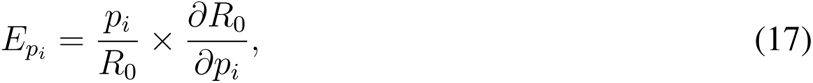

where *p_i_* is one of the parameters presented in equation 9. The local elasticities are evaluated on the fitted parameter values for Chikungunya outbreak in the mosquito model in the Austral islands, shown in Figure S3. Increasing *β* or *γ* will cause an increase in *R*_0_, while increasing *α* and *R_HM_* will cause an decrease in *R*_0_. These make intuitive sense because *β* and *γ* are the mosquito biting rate on infected people and disease transmission rate from infected mosquitoes to susceptible people, respectively. So, an increase in either one’s value is going to accelerate the spread of disease. In contrast, an increase in *α*, the rate at which the infected mosquito becomes virus-free, will cause a decrease in the average infectious period of an infected mosquito and thereby prevent the spread of the disease. An increase in the initial human-to-mosquito ratio, *R_HM_*, means a decrease in the mosquito density, the number of mosquitoes per person, which prevents the spread of the disease.

**Figure S3:**
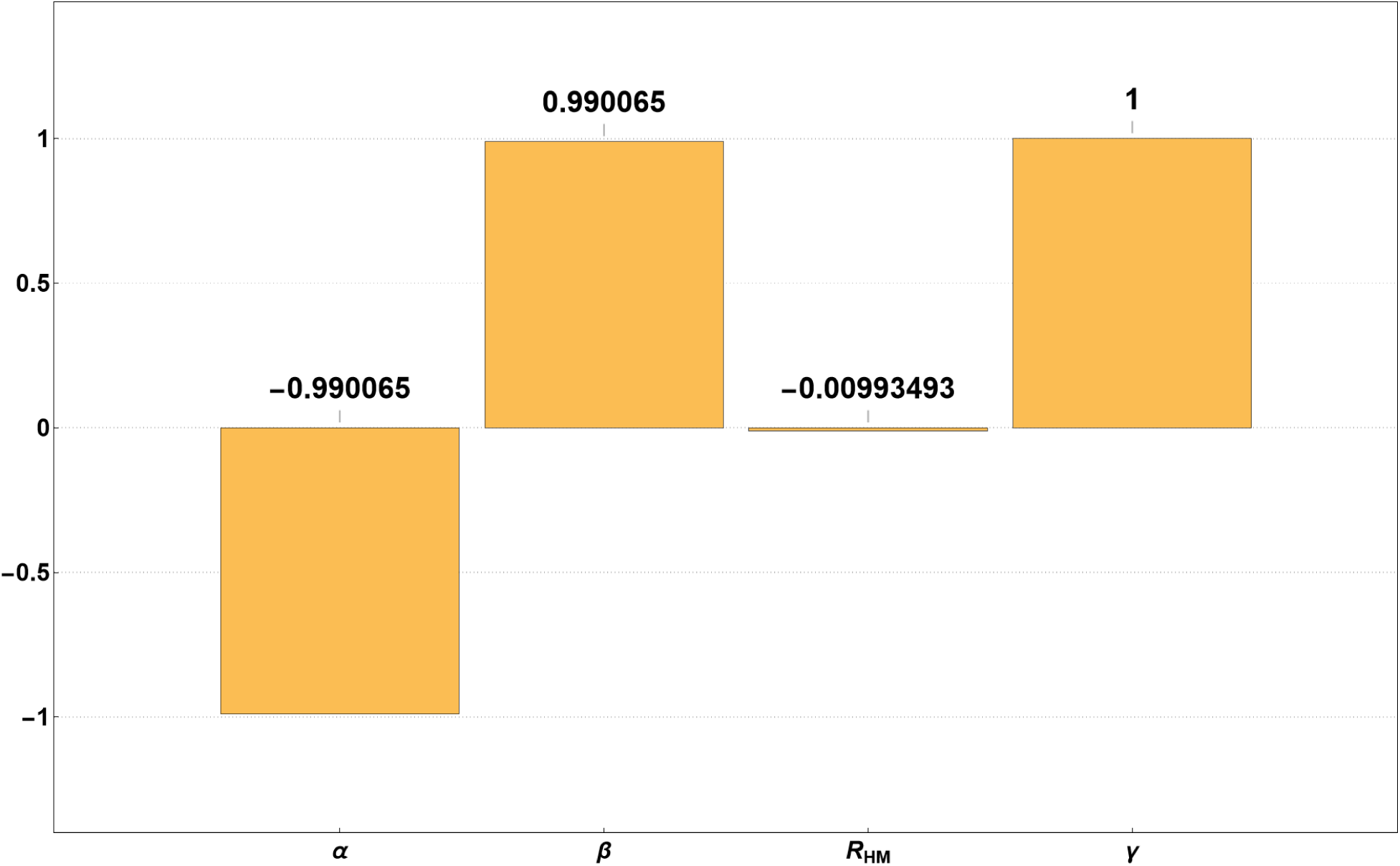
Local elasticity analysis for the mosquito model for Chikungunya outbreak in the Austral islands. The elasticity value indicates the percentage change of *R*_0_ in response to 1% increase of that parameter. Equation 17 is used to calculate the elasticity value.

## Appendix C: Model comparison using normal model

**Figure S4:**
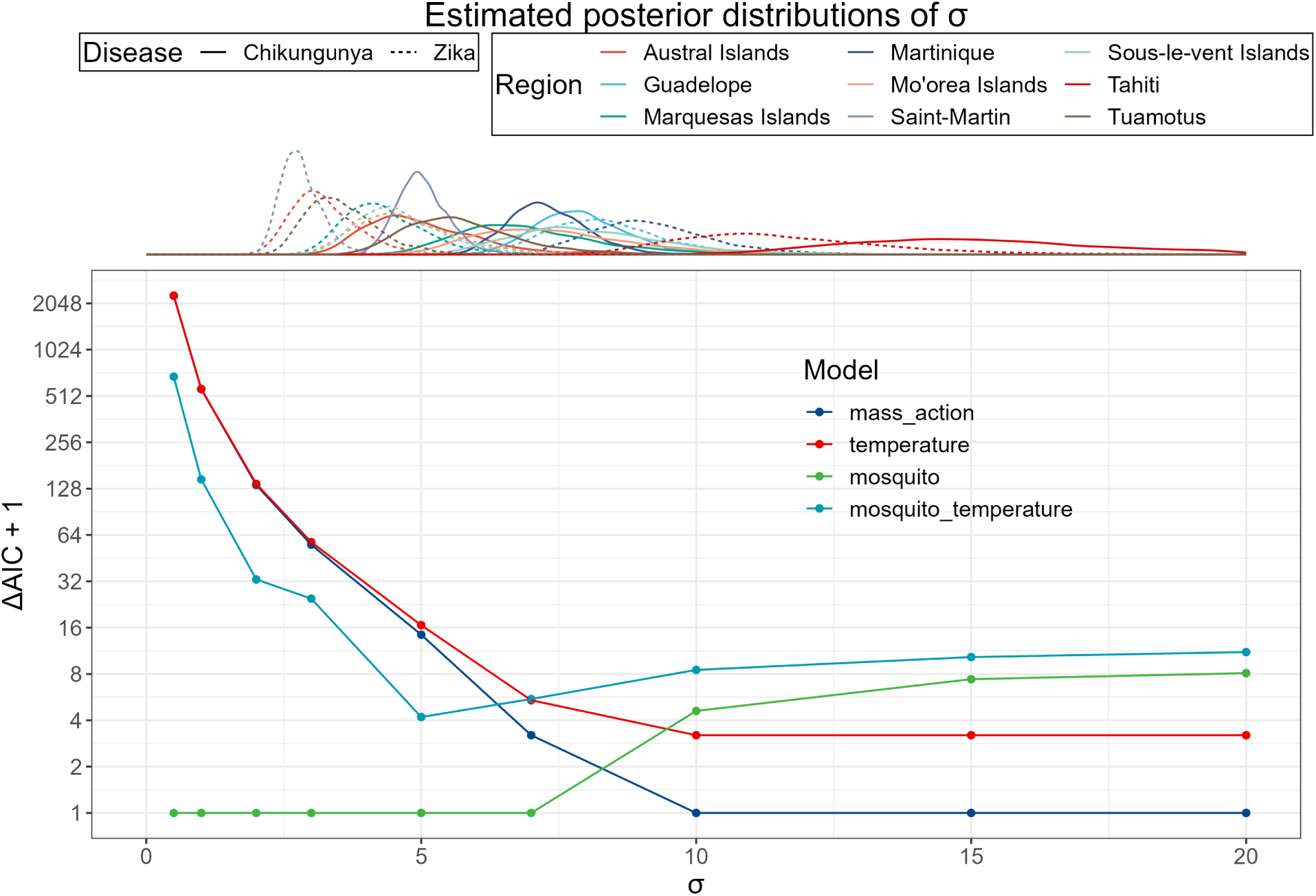
Model comparison results with Gaussian likelihood function across different values of *σ*. The top margin shows the posterior distributions of *σ* for both diseases, denoted with different line types, in 9 different regions, denoted with different colors. ΔAIC of a model is the difference of this model’s AIC and the best model’s AIC, so the model with ΔAIC + 1 = 1 is the best model. The mosquito model is the best model for *σ <* 10, and the mass action model is the best model for *σ ≥* 10. The two distributions with most of their highly probable intervals above 10 are from Tahiti, and the highly probable intervals of the other distributions of *σ* are below 10.

## Appendix D: Expected extinction time

To estimate the outbreak period, we used the mosquito model (section 2.2.2) and ignored the recovery from disease, meaning that the model is further simplified to a SI model. Then, since we only consider the disease infection, the mean first-passage time for this SI process to conclude (i.e. for the last individual to become infected) can be calculated by integrating the disease-transmission propensity with respect to the number of remaining susceptible people, as given by the equation below.

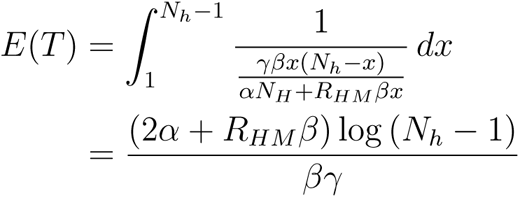

Table S1 shows the expected outbreak extinction-times of both diseases, along with the actual number of weeks that are case reported. Broadly speaking, the expected outbreak extinction-times follow the trend of the number of case-reported weeks. However, note that the actual distribution of the outbreak extinction-time is hard to parameterize due to the different population sizes in different regions, and the variance of the outbreak extinction-time will go up when the population size becomes larger.

**Table S1:** Median and 90% credible intervals of the expected number of weeks for an outbreak to end. The actual number of weeks that have reported cases are shown in the last column.

| Region | Virus | $E(T)$ (week) | Number of case-reported weeks |
| --- | --- | --- | --- |
| Austral Islands | Chikungunya | 33 [17; 126] | 18 |
|  | Zika | 32 [18; 117] | 17 |
| Guadelope | Chikungunya | 45 [25; 111] | 48 |
|  | Zika | 46 [27; 172] | 37 |
| Marquesas Islands | Chikungunya | 25 [15; 45] | 19 |
|  | Zika | 35 [19; 146] | 22 |
| Martinique | Chikungunya | 53 [26; 238] | 60 |
|  | Zika | 50 [27; 301] | 40 |
| Mo'orea Islands | Chikungunya | 33 [18; 83] | 18 |
|  | Zika | 35 [20; 149] | 16 |
| Saint-Martin | Chikungunya | 41 [21; 275] | 74 |
|  | Zika | 39 [22; 251] | 37 |
| Sous-le-vent Islands | Chikungunya | 39 [20; 123] | 21 |
|  | Zika | 37 [21; 112] | 20 |
| Tahiti | Chikungunya | 39 [23; 94] | 20 |
|  | Zika | 43 [24; 132] | 23 |
| Tuamotus | Chikungunya | 32 [18; 78] | 20 |
|  | Zika | 35 [20; 162] | 20 |

